# Comparison of the clinical characteristics and mortality in ARDS due to COVID-19 versus ARDS due to Influenza A-H1N1pdm09

**DOI:** 10.1101/2021.02.07.21251306

**Authors:** Carmen Hernández Cárdenas, Gustavo Lugo, Diana Hernández García, Rogelio Pérez-Padilla

## Abstract

**Importance:** Infection with the SARS-Cov-2 and Influenza A-H1N1 viruses is responsible for the first pandemics of the 21st century. Both of these viruses can cause severe pneumonia and acute respiratory distress syndrome (ARDS). The clinical differences and mortality associated with these two pandemic pneumonias in patients with ARDS are not well established

**Objective:** To compare case-fatality between ARDS-Covid-19 and ARDS-Influenza A (H1N1), adjusting for known prognostic risk factors.

**Design, Setting and Participants:** One hundred forty-seven patients with COVID-19 were compared with 94 with Influenza A-H1N1, all of these were admitted to the intensive care unit of the Referral Center for Respiratory Diseases and COVID-19 in Mexico City and fulfilled the criteria of ARDS.

**Results:** Patients arrived at the hospital after 9 days of symptoms. Influenza patients had more obesity, more use of Norepinephrine, and higher levels of lactic dehydrogenase and glucose, and fewer platelets and lower PaO_2_/FIO_2_. Crude mortality was higher in COVID than in influenza (39% vs. 22%; *p* = 0.005). In a Cox proportional hazard model, patients with a diagnosis of COVID-19 had a hazard ratio (HR) = 3.7; 95% Confidence Interval [CI] = 1.9-7.4, adjusted for age, gender, sequential organ failure assessment (SOFA) score, ventilatory ratio, and prone ventilation. In the fully adjusted model, the ventilatory ratio and the absence of prone-position ventilation were also predictors of mortality.

**CONCLUSION:** COVID-19 patients treated in an intensive care unit (ICU) had a 3.7 times higher risk of death than similar patients with Influenza A-H1N1, after adjusting for SOFA score and other relevant risk factors for mortality.

**Question:** Is the mortality of ARDS associated with Covid-19 greater than that of ARDS associated to influenza H1N1?

**Findings:** In a Cox proportional hazard model, patients with a diagnosis of COVID-19 had a hazard ratio (HR) = 3.7; 95% Confidence Interval [CI] = 1.9-7.4, adjusted for age, gender, sequential organ failure assessment (SOFA) score.

**Meaning:** COVID-19 patients treated in an intensive care unit (ICU) had a 3.7 times higher risk of death than similar patients with Influenza A-H1N1

## Introduction

Infection with the new Influenza A virus subtype H1N1 (Influenza A-H1N1) virus was first reported in Mexico in April 2009 (1), causing the first pandemic of the 21st century. Influenza A/H1N1 was associated with severe pneumonia and acute hypoxemic respiratory failure (AHRF) consistent with acute respiratory distress syndrome (ARDS), with an associated mortality ranging from 25.1%□41% at different sites (2). Influenza causes 390,000 deaths annually (3). The World Health Organization (WHO) estimates that influenza affect 5-10% of adults and up to 20-30% of children, especially in immunosuppressed, at extremes of life, and in persons with comorbidities(4).

The outbreak of respiratory infection by the novel coronavirus 2019 (SARS-CoV-2) started in December 2019, in Wuhan (Hubei Province), China (5, 6). From this city, the outbreak has been spreading to the majority of countries worldwide in a severe pandemic (7). Up to December 3, 2020, the COVID-19 pandemic has given rise to a total of 62 million cases and 1.4 million deaths around the world (7), mainly due to respiratory failure, although a long list of complications in and affectations to various organs and systems have been described. The COVID-19 disease is also associated with severe pneumonia, AHRF, and ARDS, requiring intensive care and ventilatory assistance in up to 5% of cases, and with a reported mortality ranging from 30-60% at different sites, with an average of 41% (8).

We considered how case-fatality compares between ARDS-Covid-19 and ARDS-Influenza A (H1N1), adjusting for known prognostic risk factors.

### Patients and setting

All patients were treated at the ICU of the National Institute of Respiratory Diseases (INER), a tertiary-level teaching and research hospital that serves the uninsured population of Mexico City and nearby states, at 2,240 meters above sea level. We obtained clinical chart information from consecutive patients with Influenza H1N1 (October 2019 to February 2020, prior to the COVID-10 outbreak) and patients with COVID-19 (March 2020 to October 2020), All of these patients had a positive RT-PCR test and fulfilled the Berlin Definition of Acute Respiratory Distress Syndrome (9). Patients with Influenza received Oseltamivir at 150 mg daily for at least 5 days, and those with COVID-19 received various treatments that frequently included corticosteroids. The study was approved by the ethics and biomedical research committee of the National Institute of Respiratory Diseases and the informed consent of the patients was dispensed because it was an observational study. The identity of the patients was protected.

Clinical and laboratory data were obtained upon admission and daily during the patients’ stay in the unit. High-resolution computed tomography (HRCT) was performed in all patients prior to admission to the respiratory intensive care unit (RICU). We calculated a bedside indicator of wasted ventilation, the ventilatory ratio (VR) as [1-minute ventilation (mL/min) × PaCO_2_ (mmHg)] /[predicted body weight x 100 x 37.5) (10) and the sequential organ failure assessment]SOFA] score (11), including PaO_2_/FIO_2_, platelets, bilirubin, creatinine, mean arterial pressure, and use of vasopressors. Glasgow score was the same as all of the patients were sedated.

The information obtained was stored in an electronic database. The survival endpoint was mortality at discharge from the ICU.

### Statistical analysis

Time to death was evaluated by a Cox proportional risk model, comparing the group with COVID-19 and that of the patients with influenza in non-adjusted models, and also adjusting for age, gender, obesity, comorbidities, and biomarkers, and mechanical ventilation parameters. Statistical analysis was performed with SPSS ver. 21 statistical software (IBM Statistics, Armonk, NY, USA) and by STATA ver. 13.0 statistical software packages. Two-sided *p* values of <0.05 were considered statistically significant.

## Results

Table 1 presents the main characteristics of patients with influenza (*n* = 94) and with COVID-19 (*n* =147). Patients with influenza had a slightly higher body mass index (BMI), lactic dehydrogenase glucose, and SOFA score, and a higher proportion of individuals with obesity, fever, shortness of breath, muscle pain, and headache, with lower neutrophils. Patients with influenza arrived for care in a worse situation (Table 1), they were more hypotense, hypoxemic, requiring vasopressors more frequently, with a higher SOFA score. Those with COVID-19 had a higher crude ICU mortality (39% vs. 22%; Fisher exact test; *p* = 0.007; log-rank test, p = 0.01), lower respiratory compliance, and spent more days in the prone position. Table 2 analyzes a multivariate Cox proportional risk model, having as a dependent variable time to death in the ICU and, as an independent variable, the type of virus (COVID-19 vs influenza) adjusted by age, sex, comorbidities, symptoms, laboratory results, and ventilatory parameters: COVID-19 patients had nearly 4 times more risk for death (OR = 3.7; 95% CI = 1.9-7.4) than patients with influenza. Additional variables associated with predicting mortality were the ventilatory ratio, the SOFA score, and th the BMI, whereas prone ventilation was associated with reduced mortality (see Table).

**Table 1.**
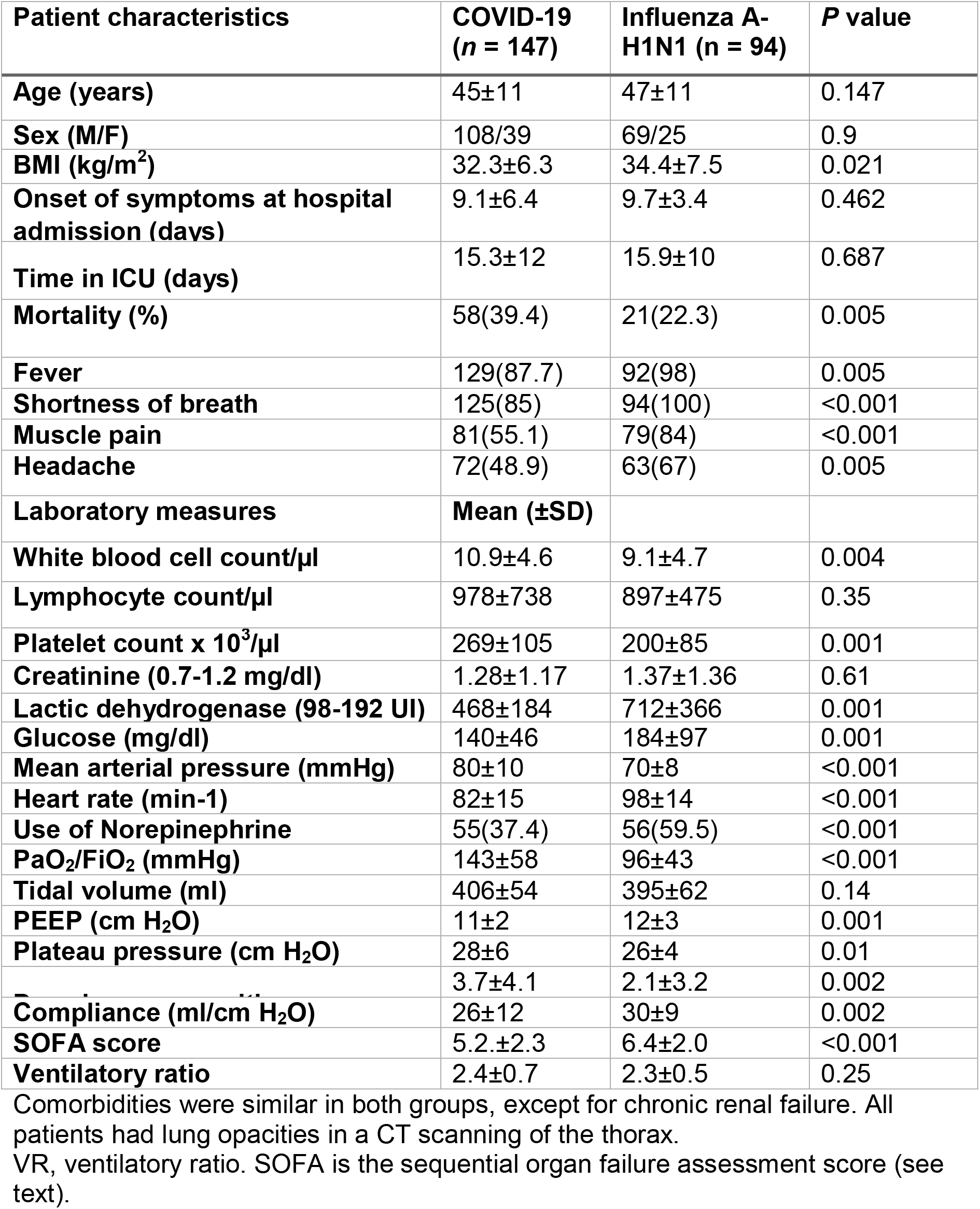
Demographic and clinical biomarkers and radiologic characteristics of the patients.

**Table 2.** Adjusted multivariate Cox proportional hazard models.

|  | Adjusted 1 |  | Adjusted 2 |  |
| --- | --- | --- | --- | --- |
| HR | HR (95% CI) | *P* | HR (95% CI) | *P* |
| COVID-19 | 5.1 (2.7-9.7) | <0.001 | 3.7 (1.9-7.4) | <0.001 |
| SOFA score | 1.5 (1.4-1.7) | <0.001 | 1.3 (1.2-1.5) | <0.001 |
| Glucose (mg/dl) | 1.005 (1.002-1.008) | 0.002 | 1.0 (1.0-1.01) | 0.02 |
| BMI (kg/m2) | 1.02 (0.99-1.05) | 0.13 | 1.03(1.01-1.06) | 0.03 |
| Days in prone position |  |  | 0.94 (0.88-1.001) | 0.055 |
| Ventilatory ratio |  |  | 2.3 (1.5-3.4) | <0.001 |
| Akaike information criterion | 679 |  | 649 |  |
| Bayesian information criterion | 693 |  | 670 |  |
| Log likelihood of model | -335 |  | -318 |  |

BMI, body mass index; VR, ventilatory ratio; HR, Hazard Ratio. SOFA is the sequential organ failure assessment score (see text). For the crude model (only COVID-19). Log likelihood -371, AIC 744, and BIC 748.

## DISCUSSION

Patients with COVID-19 had a substantially higher lethality than those with influenza, despite both being in respiratory failure, both with ARDS, and both after taking into account various predictors of death such as age, sex, some laboratory measurements, and ventilatory parameters, indicating in COVID-19 more severe lung damage and fewer possibilities of recovery.

Influenza and COVID-19 may cause severe damage to the lungs, to other organs, and may cause death, and it is unknown whether this may be attributed to direct viral injury, to an exaggerated inflammatory reaction, or to the often mentioned cytokine storm (12, 13) in addition to personal susceptibility. Patients with COVID-19 have been compared with those with influenza and lacked a difference in cytokine levels (13). The findings of CT scanning overlap in the two infections (14), but patients with COVID-19 manifested vascular inflammation, and immunothrombosis (15) to a higher degree than those with influenza. In addition, there is an antiviral treatment for influenza (all of our patients received this treatment), and a vaccine of variable affectivity is available, although only a small proportion of our patients are vaccinated (5% of those arriving at the ICU). The majority of our patients with COVID-19 received systemic corticosteroids, but an antiviral treatment and a vaccine were not yet available.

Our study shows limitations such as a small cohort of patients and coming from a single high specialty center. However, it is prospective, and the cohorts were contemporary.

In conclusion, patients with respiratory failure and ARDS from COVID-19 treated in an ICU entertain a substantially increased risk of death compared with similar patients with Influenza A-H1N1, even adjusting for SOFA score and other relevant risk factors for mortality. Differences in their pathogenic mechanisms and the absence of antiviral drugs and vaccines likely contributes to worse outcome.

**Figure 1.**
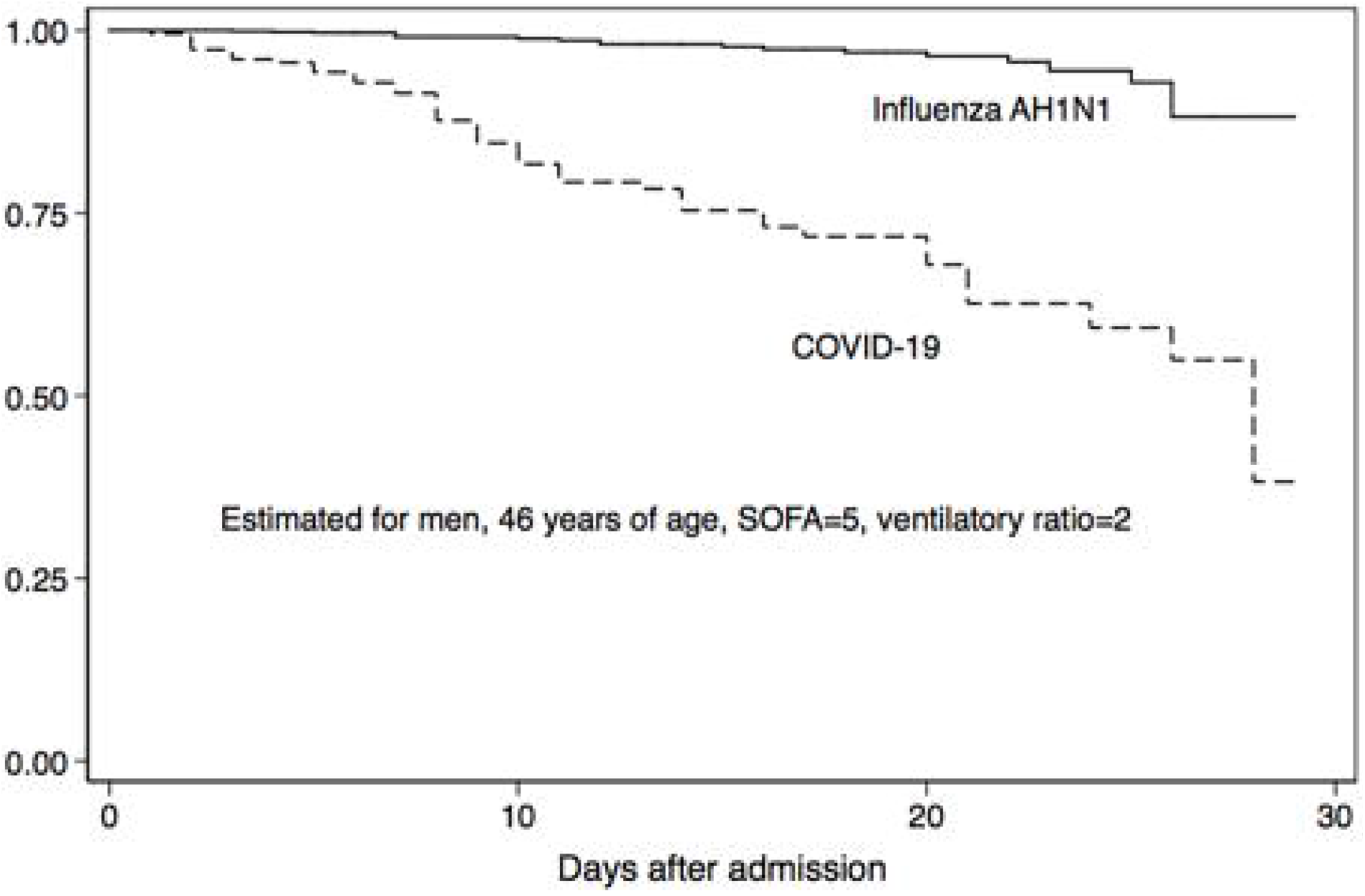
Kaplan-Meier survival curve for COVID-19 patients (dashed line) and for patients with Influenza A-H1N1 (continuous line) adjusted for sex, age, sequential organ failure assessment (SOFA) score (SOFA = 5), and ventilatory ratio (VR = 2).

## Data Availability

the data are available to interested researchers upon justification and request to the authors.

